# Novel indicators for evaluating topological threats to populations from pandemics applied to COVID-19

**DOI:** 10.1101/2020.05.29.20116491

**Authors:** Shun Adachi

## Abstract

To deal with pandemics, evaluating the temporal status of an outbreak is important. However, prevailing standards in this respect are mostly empirical and arbitrary. As an alternative, we focus on a novel approach which configures indicators that evaluate topological threats to populations due to the COVID-19 pandemic. We extended the current PzDom model to calculate a threshold of the model for accelerated growth, an indicator of growth extent Re(*v*), covariance Re(*s*), a topological number *E*(*l*), and expected sums of possibly increasing numbers of infected people. We term this the exPzDom model. The indicators in the exPzDom model adhere well to the empirical dynamics of SARS-CoV-2 infected people and align appropriately with actual policies instituted by the Japanese government. The described indicators could be leveraged pursuant of objective evaluation based on mathematics. Further testing of the reliability and robustness of exPzDom model in other pandemic contexts is warranted.

## Introduction

To understand and respond appropriately to pandemics, it is important to evaluate the temporal status of a disease outbreak. However, standards in this case are often empirical and arbitrary. It is difficult to apply SIR and related models in some cases, particularly where a given pandemic is not significantly influenced by herd immunity amongst the population [e.g., Koroveinikov and Maini (2004), Gumel et al. (2004), Longini et al.(2004)]. For example, a small decrease in new infections following a sharp increase or flattened peak beforehand, as has been frequently observed in SARS-CoV-2 infection dynamics, is difficult to explain by a simple model. Therefore, an alternative approach that tackles the issue qualitatively with topological analysis might have important merits. Our aim here is to put forward a novel approach for indicator development that evaluates topological threats to populations from COVID-19 in Japan and the rest of the world.

## Methods

Data were taken on new cases of SARS-CoV-2 infections validated by PCR from 3/12/2020 to 10/20/2020 in 47 prefectures in Japan (https://github.com/kaz-ogiwara/covid19) and from 2/24/2020 to 10/20/2020 in 20 highly affected countries around the world (https://github.com/CSSEGISandData/COVID-19). For Japan, serious cases for COVID-19 were also analyzed from 3/24/2020 to 10/20/2020 in 47 prefectures in Japan. The PzDom model, proposed in Adachi (2019a), contains a set of indicators to analyze population dynamics. It describes a real part of a complex metric Re(*s*) as an indicator of divergence beyond Gaussian fluctuation. Re(*s*) is calculated as follows. First, the group is sorted by the rank *k* of population numbers *N*_*k*_s. If *k* ≠ 1, Re(*s*) = ln(*N*_*1*_/*N*_*k*_)/ln *k*. For *k* = 1, this indicator is calculated by an inverse Riemann ζ function, regarding ζ(*s*) = ∑*N*_*k*_/*N*_*1*_. For replacing an imaginary part of the metric Im(*s*) = exp(∑*N*_*k*_*Re(*s*)/*b*), where *b* is an approximated parameter of a distribution *N*_*k*_ = *a* – *b* ln *k* at a particular point in time, a new Im(*s*) is calculated by (∑*N*)*Im(*s*)^^1/”Number of data groups”^, where ∑*N* is the sum of all infected people at a particular time point. This modification is instituted because in the PzDom model, the purpose of Im(*s*) is to predict a maximum for the growth of a particular biological species. Herein, we would like to focus on an expected sum of all newly infected people among the examined regions with Im(*s*) as “expected sums”. Next, Re(*v*) is developed in Adachi (2017) and Adachi (2019b). Re(*v*) = ln *N*_*k*_/ln(Im(*s*)) and it is an indicator of non-Archimedean valuation of *N*_*k*_. It represents how far an observed group has a tendency for potential growth. *E*(*l*) is proposed in Adachi (2019b) and it represents a categorical value of the observed set of groups, converged to a certain value. That is, a number of external symmetries of observed groups. Note that the convergence value is a function of the applied dataset and this value would be different between empirical applications even where data overlap exists. It is calculable by Im(*v*) = exp(∑*N*_*k*_*Re(*v*)/(“Number of data groups” * *b*)) (this time focusing on individuals, not whole sum), *E*(*l*) = ln *N*_*k*_/ln Im(*v*). The threshold is proposed in Adachi (2019c) as a combination of blackhole-like analogy of a growth speed *D* = exp(Re(*s*)/*b*) from the PzDom model (Adachi 2019a) and a Jeans wavelength-like parameter λ*J* = 2π*sqrt(*bD*/*N*_*k*_Im(*s*)) (Adachi 2019c). The threshold is *D*/λ*J* = 1 at a unit time and if the value exceeds 1, growth would be accelerated analogical to escaping from a particular black hole when an object is not within the Schwarzschild radius. We refer to this extended version of the PzDom model as “exPzDom”. The reports of new cases involve systematic differences in PCR analyses, and sometimes the values are NaN or negatives (errors for reports), which are regarded as zeros in our model. Code in Julia language describing this new model is publicly available via the following link: https://github.com/sadachi79/exPzDom.

## Results

We performed calculations of the indicators described above and obtained the following results. Raw data are shown in Fig. 1.

**Fig. 1.**
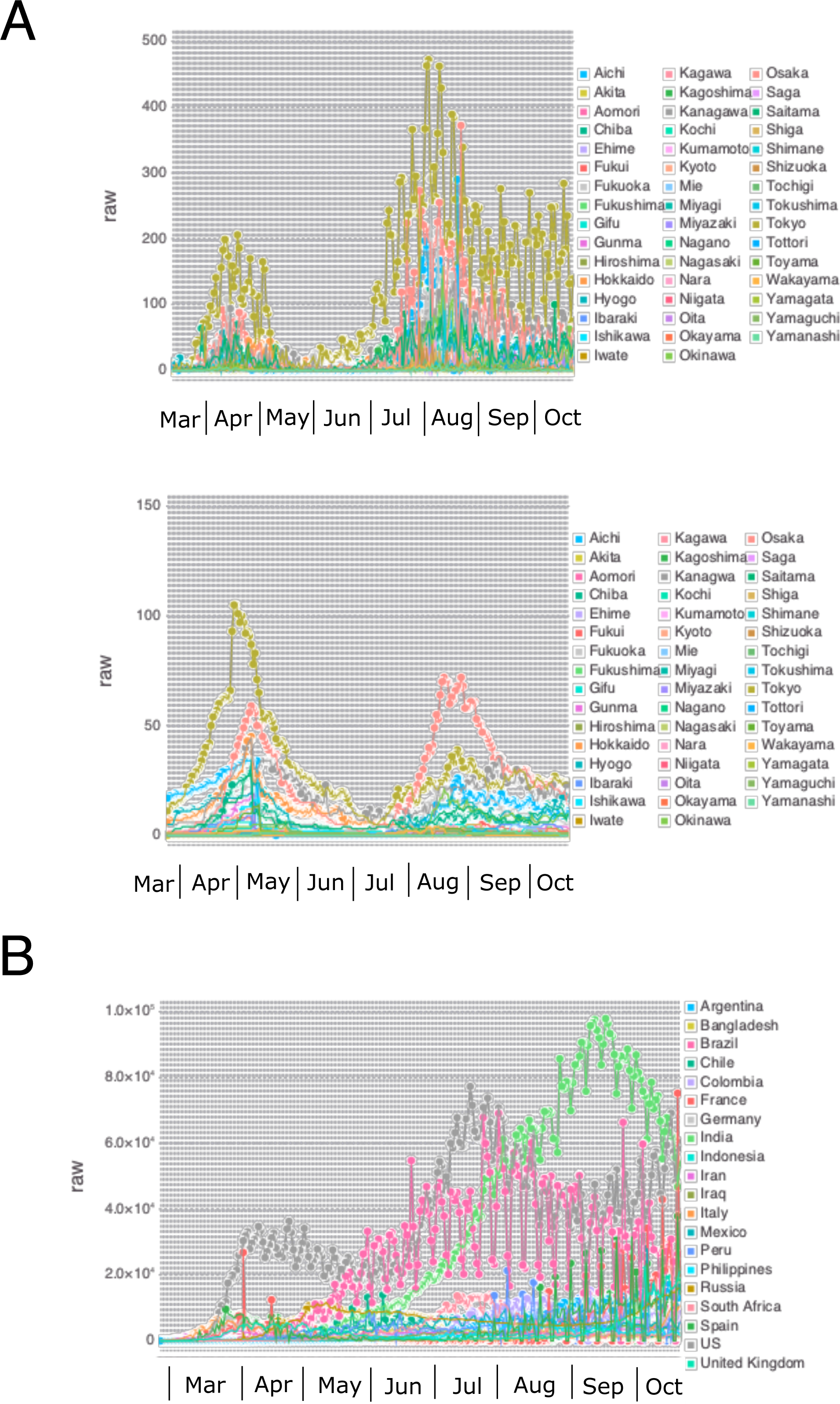
Raw values for newly infected and serious cases. **A**, Japan. Top, PCR. Bottom, serious cases for COVID-19. **B**, World.

There was a significant increase in the “threshold” of PCR results in Japan until mid-April. Approximately 2 weeks after the declaration of the state of emergency and partial lockdown by the Japanese government on April 7, 2020, the threshold value started to decrease and this well represents the decrease in new cases during this period. Serious cases for COVID-19 followed slightly after the results for PCR (Fig. 2A). Tokyo was prominent in dynamic behavior (Fig. 2A). Comparing with raw results, threshold can easily remark the increase of data from end-May to August, which is difficult to recognize in raw data before July (Figs. 1A and 2A). In terms of the global situation, the poor situation of the U.S.A., Brazil and India are obvious, with diverging growth of COVID-19, followed by other countries. The leading countries shifted from United Kingdom, Italy, U.S.A., Brazil, U.S.A. to India (Fig. 2B).

**Fig. 2.**
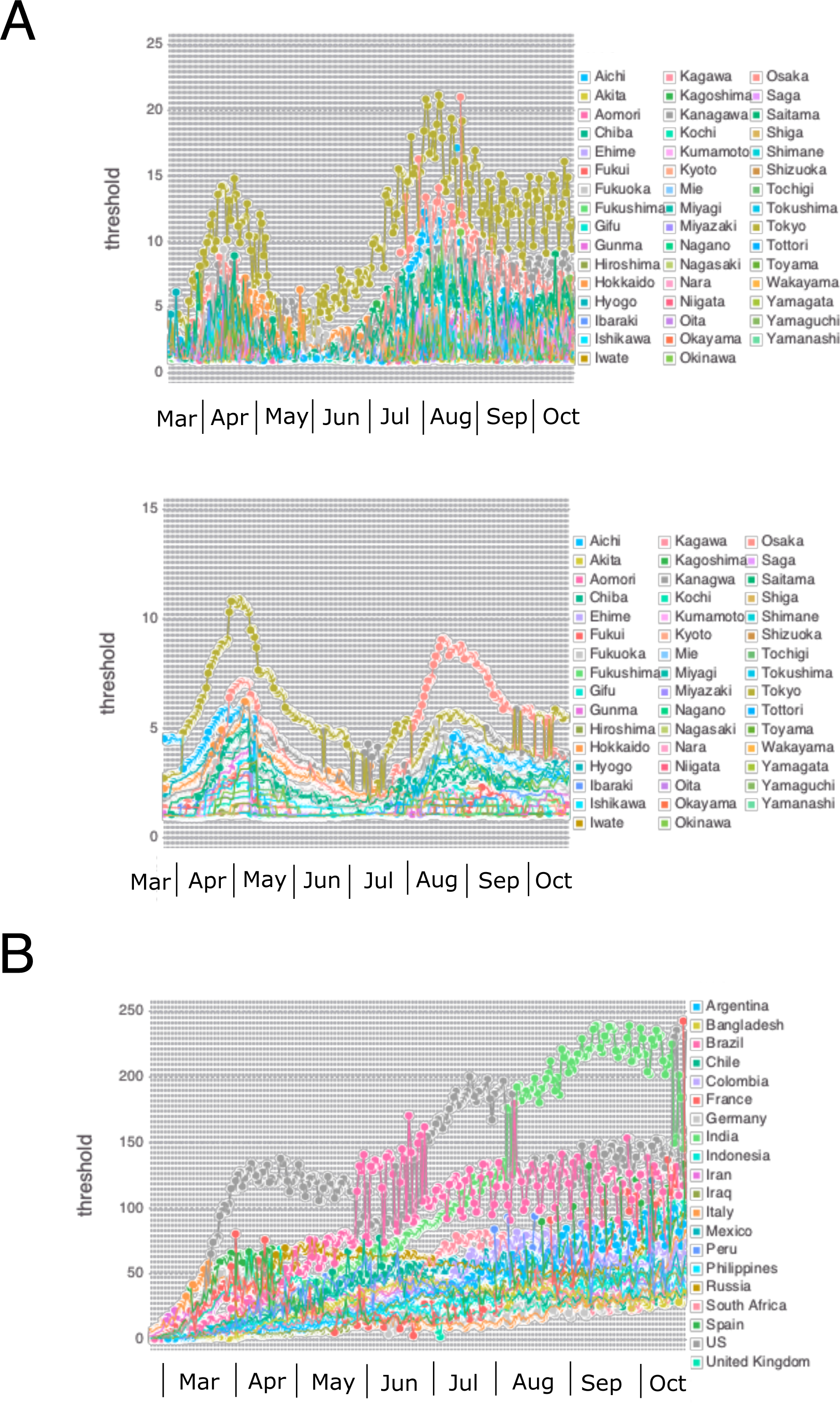
Threshold values for exPzDom model. **A**, Japan. Top, PCR. Bottom, serious cases for COVID-19. **B**, World.

Re(*v*) in Japan can evaluate qualitative differences among the 13 prefectures subjected to special cautions defined by the Japanese government (from April 16, 2020 to May 14, 2020; probable infections with the disease were supposed to be 2 weeks before this, possibly during April 2020) and the remaining 34 prefectures. The former ranged mostly between 0.3 and 0.8 (disregarding Ibaraki and Gifu with lower values), while the latter ranged mostly between 0.1 and 0.4 during April. Similar things were observed in the second wave peaked in August (Fig. 3A). Serious cases for COVID-19 followed slightly after the results for PCR (Fig. 3A). For the world, again the U.S.A., Brazil and India are notable with diverging growth of COVID-19. The other countries follow this trend to a lesser extent. The burst during March is easily recognizable (Fig. 3B).

**Fig. 3.**
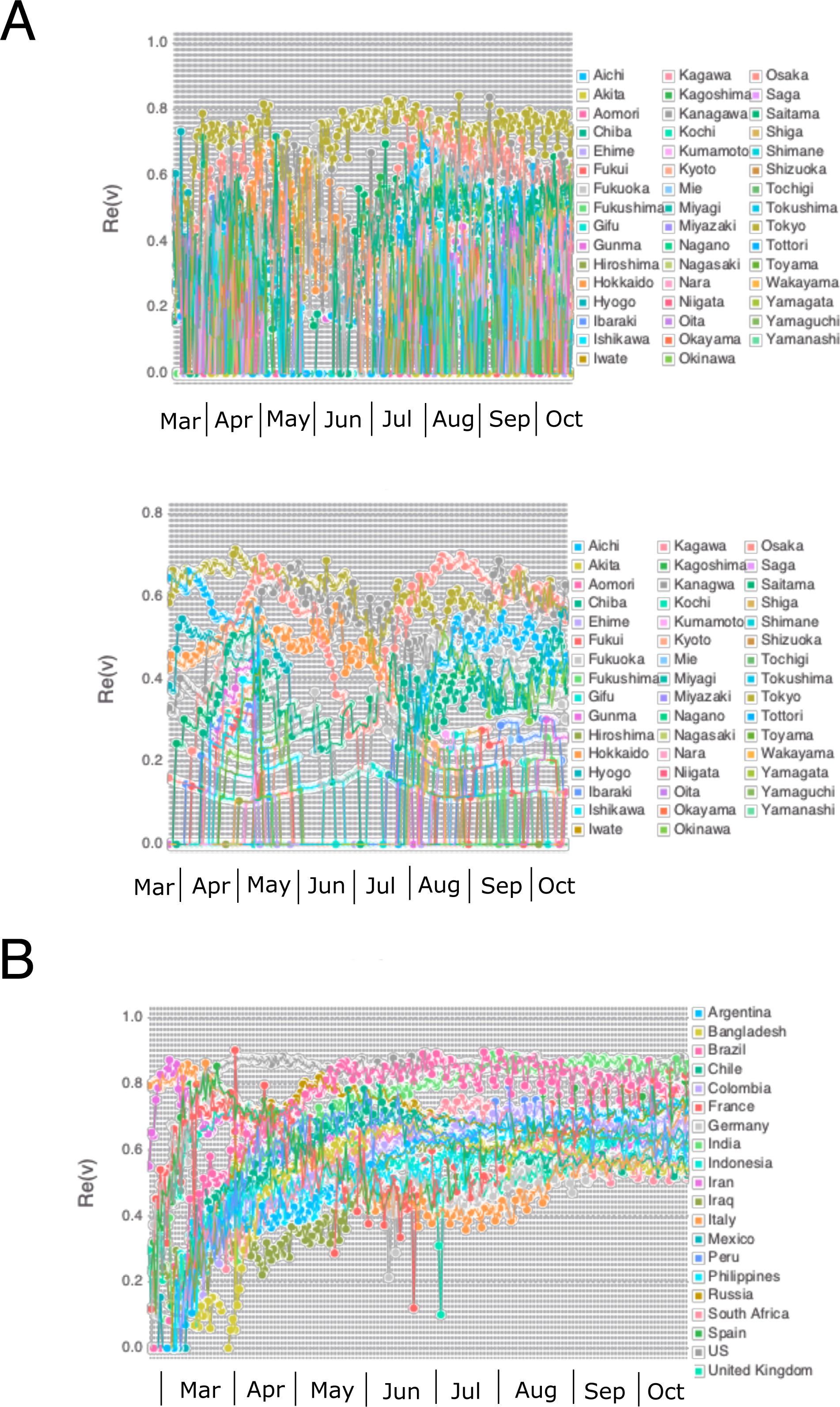
Re(*v*) values for exPzDom model. **A**, Japan. Top, PCR. Bottom, serious cases for COVID-19. **B**, World.

Re(*s*) is an indicator related to the covariance of the data, and 0 < Re(s) < 1 means the extent of acceleration in the population is decreasing, while 1 < Re(s) < 2 means increasing, within the category of Gaussian fluctuations. Re(*s*) = 1 is a neutral situation. Re(*s*) > 2 means an explosive increase/decrease. Further information is available in Adachi (2019a). For Re(*s*) in Japan, infections were sporadic until March 22, 2020; the growth acceleration mode then shifted (in this case, increased) until May 6, 2020. Subsequent to this, the mode shifted back to a rather repressive mode. However, it again shifted to acceleration from end-May to June and shifted again to repressive mode in July. After August, the values were more or less stable. These interpretations well represent the actual dynamics in the growth of infected people. It can predict shifts of the mode before they are easily recognizable by raw data (Figs. 1A and 4A). Serious cases for COVID-19 followed after the results for PCR (Fig. 4A). For the world, explosions with Re(*s*) > 2 ended in end-March. Explosions in individual countries such as France and United Kingdom were observed during June. Basically most of the countries restricted to 1< Re(*s*) < 2 mode as moderate growth mode, while some are close to Re(*s*) = 2 (previously Italy, France and Iraq were) and some below Re(*s*) = 1 with retardation mode (India and Brazil, in recent days, while for some periods U.S.A. was) (Fig. 4B).

**Fig. 4.**
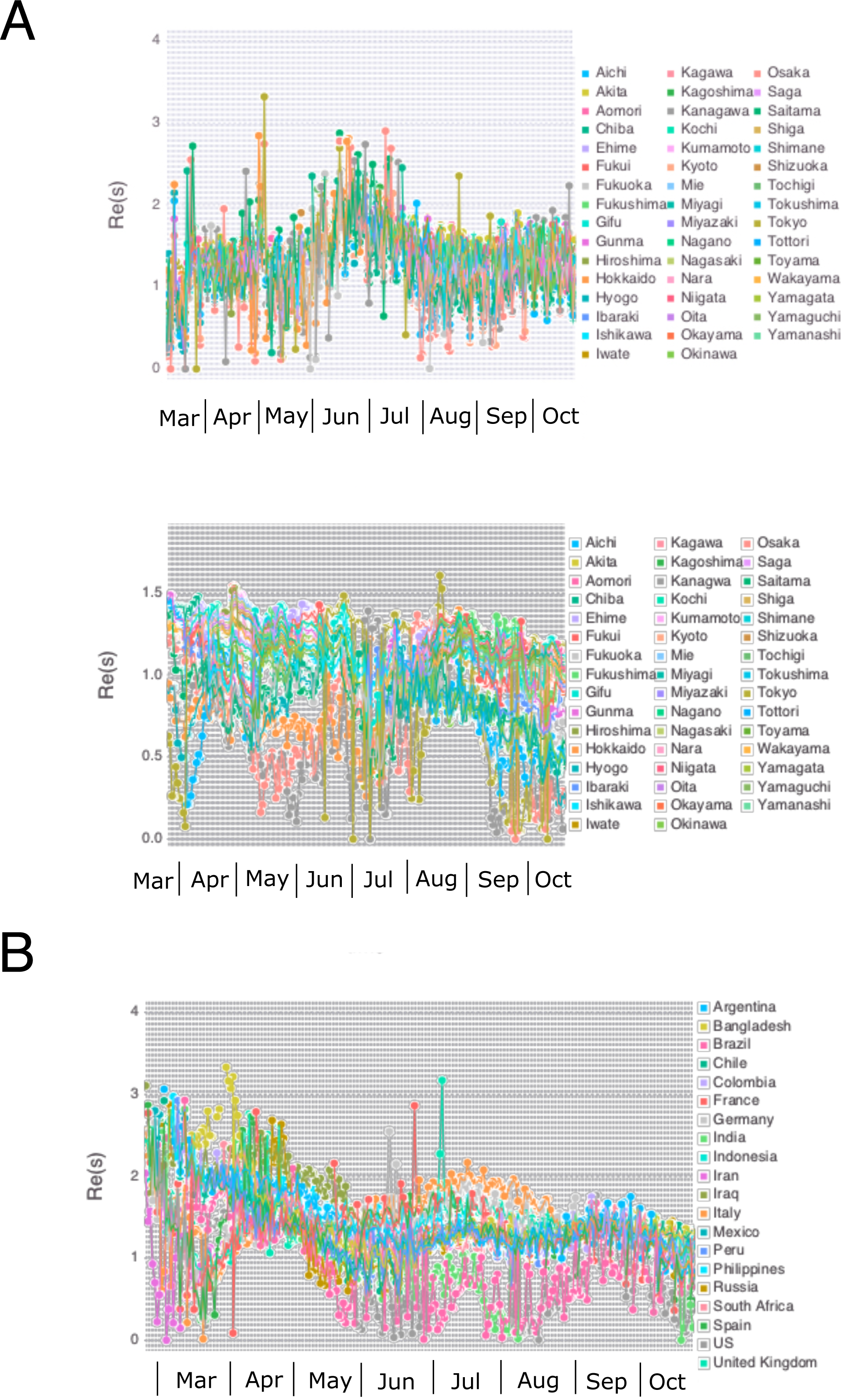
Re(*s*) values for exPzDom model. **A**, Japan. Top, PCR. Bottom, serious cases for COVID-19. **B**, World.

Moving on to *E*(*l*), in Japan, empirically *E*(*l*) ∼ 10 until March 24, 2020 was a safer course; *E*(*l*) ∼ 15 during April 2020 was problematic; and *E*(*l*) ∼ 20 at the beginning of May 2020 represents danger, followed by a significant decrease to *E*(*l*) ∼ 10 again. It started to increase again till July, reaching ∼20 and then slightly decreased to 15 or so. It predicts the actual dynamics of raw data (Fig. 1A and 5A). Serious cases for COVID-19 followed after the results for PCR (Fig. 5A). For the world, the top 20 countries still seem to be in a dire situation severer than Japan. It was retarded from April to May, however, again the values went up since then. After October, the level came down to ∼20 (Fig. 5B). Note that *E*(*l*) would converge to a particular value at each time point, regardless of overlapping sets of data. If individual datums are somewhat different, it would converge to a different value.

**Fig. 5.**
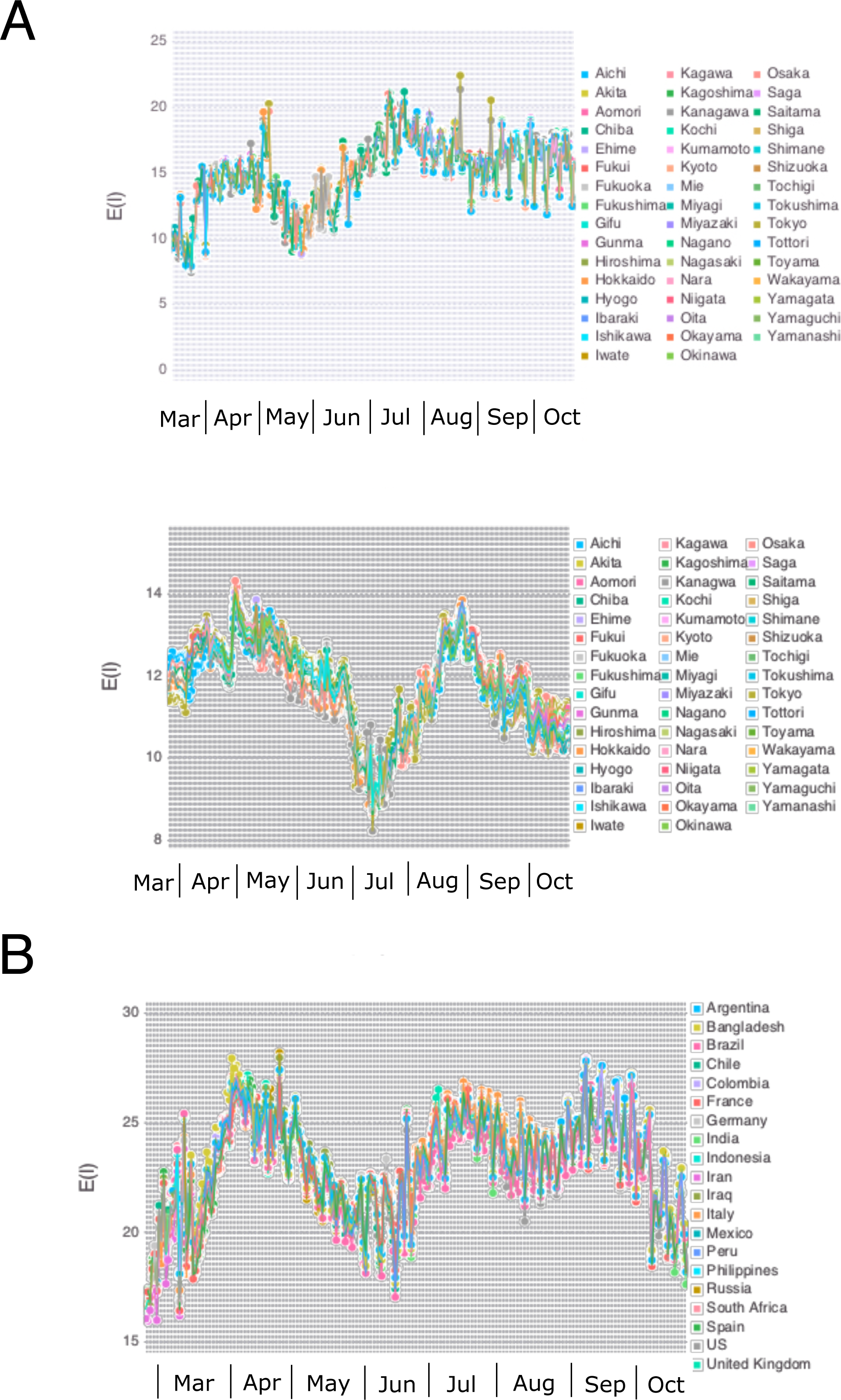
*E*(*l*) values for exPzDom model. **A**, Japan. Top, PCR. Bottom, serious cases for COVID-19. **B**, World.

For “expected sums” in Japan, it seems that hundreds of people are expected to be infected (Fig. 6A). Serious cases for COVID-19 followed after the results for PCR (Fig. 6A). For the world, it appears that hundreds of thousands of people still have potentials for infection every day. After explosion until March and succeeding phase of constant values in April and May, moderate growths from June occurred till July. August and September were in more or less stable mode. From October, either acceleration or repression was observed, different from country to country (Fig. 6B).

**Fig. 6.**
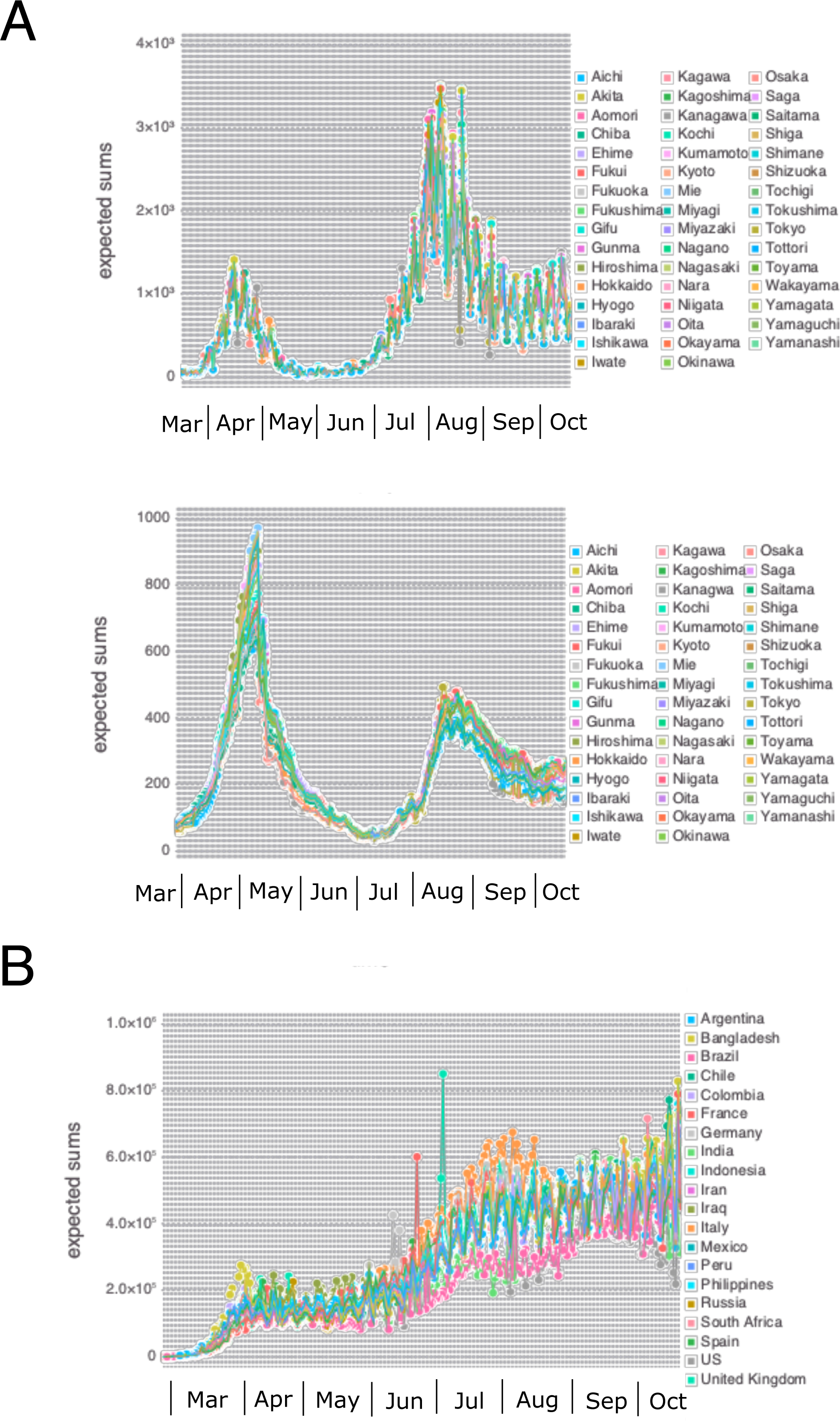
Expected sums for exPzDom model. **A**, Japan. Top, PCR. Bottom, serious cases for COVID-19. **B**, World.

## Discussion

Through the newly developed exPzDom model, we can calculate threshold, Re(*s*), Re(*v*), E(*l*), and expected sums. As shown, these indicators provided a good fit to the actual development of policies by national governments and could therefore be usefully leveraged in decision-making contexts pursuant of objective evaluation based on mathematics. The threshold indicator captures the dynamics of explosive dangers, and Re(*v*), Re(*s*), and E(*l*) are topological indicators for the accelerative growth of infections among populations. Expected sums, although they fluctuated markedly, might indicate the worst-case scenario deduced according to available data from the current point in time. A limitation of this study is that it utilizes new cases of infection as data, not actual onsets of the disease, which would reflect biological dynamics better. Further exploration of the reliability and robustness of the exPzDom model using data from other pandemics is warranted to better develop objective indicators which can be effectively used in decision-making contexts.

## Data Availability

All the data and a code for the analyses are available via indicated links.

https://github.com/kaz-ogiwara/covid19

https://github.com/CSSEGISandData/COVID-19

https://github.com/sadachi79/exPzDom

## Declarations

### Funding

There was no funding for this work.

### Conflict of interest/competing interests

The authors declare that they have no conflict of interest.

### Ethical approval

N/A.

### Consent to participate

N/A.

### Consent for publication

N/A.

### Availability of data and material

All the data are available from https://github.com/kaz-ogiwara/covid19 and https://github.com/CSSEGISandData/COVID-19.

### Code availability

The code is available from https://github.com/sadachi79/exPzDom.

### Authors’ contributions

SA designed, coordinated the study and ran the literature search. All authors acquired data, screened records, and extracted data. SA did mathematical analyses and wrote report. All authors provided critical conceptual input, analyzed and interpreted data, and critically revised the report.

## Acknowledgments

I am very grateful to reviewers and colleagues for their comments and advice spanning medicine, biology, mathematics, and physics. The findings and conclusions in this report represent the analysis and interpretations of the author and do not necessarily represent the official position of Kansai Medical University.

## Notes

### Competing Interest Statement

The authors have declared no competing interest.

### Funding Statement

There was no funding for this project.

### Author Declarations

IRB/oversight body is exempted.

### Summary of Updates

The data were refined with new descriptions. The errors for equations were also corrected.

